# A technology-based randomized controlled trial of self-affirmation and gain-framed health messaging to reduce sedentary behavior in older adults

**DOI:** 10.1101/2025.02.18.25322482

**Authors:** Meishan Ai, Nagashree Thovinakere, Caitlin S. Walker, Cora Ordway, Elizabeth Quinoñez, Emily Melsky, Frank D’Agostino, Calvin Tobias, Susan Whitfield-Gabrieli, Siobhan M. Phillips, Dominika M. Pindus, Charles Hilman, Timothy P. Morris, Arthur F. Kramer, Maiya R. Geddes

**Author notes:** These authors contributed equally to this work (Meishan Ai, Nagashree Thovinakere, Caitlin S. Walker). Corresponding Author: Maiya Geddes.

## Abstract

**Objective:** Sedentary behavior significantly increases the risk for chronic diseases and cognitive decline in aging, underscoring the need for effective interventions. Older adults exhibit a ‘positivity effect’, whereby processing of positive information is prioritized over negative information. In addition, self-affirmation was shown to reduce sedentary behavior in younger adults, but its effects in older adults remain unknown. This study tested a novel, technology-based intervention combining daily self-affirmation and gain-framed health messages to reduce sedentary behavior in older adults.

**Methods:** In a 6-week randomized controlled trial (NCT0431536), 48 cognitively unimpaired, sedentary older adults were randomized into two groups: The intervention group (mean age=70.0±5.4years) completed daily self-affirmation based on their highest-ranked value, followed by gain-framed health messages. The active control group (mean age=68.4±5.0years) performed self-affirmation on their lowest-ranked value, followed by loss-framed messages. This was a single-blinded clinical trial that incorporated a hybrid efficacy and implementation design. Thus, information about intervention feasibility was examined. In addition, baseline motivational traits, including reward sensitivity, were assessed as moderators of behavior change. The neural basis of self-affirmation and gain-framed health messaging was examined at baseline using a task-based, event-related fMRI paradigm across groups, after randomization at the outset of the intervention.

**Results:** The intervention showed high adherence (0.92±0.08) and positive ease-of-use ratings. While the intervention did not significantly reduce sedentary behavior compared to the active control condition, increased reward sensitivity predicted reduced sedentary behavior across all participants. FMRI results showed increased ventral striatal activation in the intervention group, compared to the active control group during reading of gain-framed compared to neutral messages.

**Conclusions:** This study supports the feasibility of technology-based sedentary beahvior reduction interventions for older adults. While self-affirmation combined with gain-framed messaging did not significantly reduce sedentary behavior, gain-framed messages engaged the reward network, and reward sensitivity predicted future reduction in sedentary behavior.

## Introduction

Sedentary behavior is highly prevalent among older adults, with 67% of the population spending over 8.5 hours daily engaged in sedentary activities (Mathews et al., 2019). This problem has been exacerbated by the COVID-19 pandemic, which significantly increased sedentary time among older adults (Oliveira et al., 2022). Excessive sedentary behavior, defined as wakeful physical behaviors performed while sitting, lying or reclining, with energy expenditure ≤ 1.5 metabolic equivalents, is associated with elevated risks of all-cause mortality, cardiovascular disease, cancer, Type 2 diabetes, and cognitive decline (Bailey et al., 2019; Ekelund et al., 2019). One metabolic equivalent represents the amount of oxygen consumed while sitting at rest (3.5 ml O₂ per kg body weight per minute) (Ekelund et al., 2019). Additionally, sedentary behavior imposes a significant economic burden on healthcare systems: In Canada for example, it accounts for an estimated $2.2 billion annually, with potential savings of $219 million achievable through a 10% reduction in sedentary time among adults (Chaput et al., 2022). Notably, sedentary behavior is distinct from, and has dissociable health effects compared to, insufficient physical activity (Spence et al., 2017; van der Ploeg & Hillsdon, 2017). For example, a person can both perform 30 min of moderate-to-vigorous physical activity achieving recommended levels and also engage in a high volume of sedentary behavior throughout the rest of the day.

How to ideally motivate and support sedentary behavior reduction is a critical unanswered question. The socioemotional selectivity theory posits that aging is accompanied by shifts in motivation, whereby older adults prioritize social emotional well-being compared to younger adults (Charles, Mather, & Carstensen, 2003). Interventions that change social context for example has been shown to be effective at reducing sedentary behavior (Gardner et al., 2016). Older adults exhibit a ‘positivity effect’, wherein they preferentially attend to and remember positive compared to negative information (Knutson et al., 2001; Charles, Mather, & Carstensen, 2003). For example, when shown positive, negative, and neutral images, older adults demonstrate better recall for positive compared to negative images, to a greater degree than younger adults (Charles, Mather, & Carstensen, 2003). This effect extends to the influence of health messaging on physical activity engagment: Gain-framed health messages that emphasize the benefits of physical activity were shown to enhance step count in older adults (Notthoff and Carstensen, 2015). A prior study demonstrated that older adults exposed to positive messages about the benefits of walking increased their walking behavior significantly more than those exposed to risk-based or neutral messages. Given that subtle differences in message framing can influence health behavior, testing the impact of gain-framed messages on sedentary behavior in older adults is a promising potential strategy for behavior change.

One significant challenge in reducing sedentary behaviors through health messaging, however, is the perception of self-relevant health messages as threatening to self-worth, often resulting in resistance to change. This issue highlights a paradox, where individuals at the highest risk are often the most defensive, decreasing their receptivity to altering risk behaviors (Cohen & Sherman, 2014). Moreover, a single strategy is less effective in reducing sedentary behaviors than leveraging multiple behavior change techniques. Self-affirmation, a psychological technique involving reflection on core values, was shown to mitigate defensiveness and enhance openness to behavior change (Falk et al., 2015). By affirming their values before encountering potentially threatening health messages, individuals bolster their self-integrity, thereby reducing resistance and increasing the likelihood of behavior change (Cooke et al., 2014; Falk et al., 2015). Among younger adults, the efficacy of self-affirmation was demonstrated across various health domains, including smoking, willingness to undergo diabetes screening, and acohol consumption behavior (Kang et al., 2018). In the domain of physical activity, prior combined behavioral and functional magnetic resonance imaging (fMRI) research has shown that younger individuals that reported high self-affrmation exhibit greater reductions in sedentary behavior compared to individuals who report low self-affirmation(Sweeney & Moyer, 2015). Taken together, this evidence suggests that combining gain-framed health messaging with self-affirmation may be particularly effective at reducing sedentary behavior among older adults. The present study extends this body of research by evaluating the efficacy of an intervention that combines gain-framed health messaging and self-affirmation to reduce sedentary behavior in older adults, addressing a critical gap in understanding strategies for promoting active aging.

Understanding the behavioral and neural mechanisms underlying changes related to sedentary behavior is critical for advancing theoretical models of behavior change and informing the development of theoretically-motivated evidence-based interventions. Behavioral and neuroimaging studies support the relevance of reward-related processes in sedentary behavior change (Falk et al., 2010, 2015; Morris et al., 2022). Reductions in sedentary behaviour have been shown to be driven by reward sensitivity, including the anticipated pleasure associated with the adoption of healthy lifestyles (Michaelsen & Esch, 2023). Neuroimaging studies in younger and midlife adults have shown that self-affirmation activates key brain regions involved in self-referential processing, such as the ventromedial prefrontal cortex (vmPFC), and reward valuation, such as the ventral striatum (VS) (Falk et al., 2015; Morris et al., 2022). Activation in these regions during an event-related fMRI task at baseline, was associated with significant future reductions in sedentary behavior during a four-week self-affirmation intervention in younger adults (Falk et al., 2015; Casio et al., 2016). The vmPFC and VS are nodes in intersecting circuits involved in reward valuation and self-referential processing (Casio et al., 2016; Bartra et al., 2013). These regions have also been implicated in the processing of persuasive messages. For instance, individual differences in vmPFC activity during exposure to persuasive health messages have been shown to predict changes in sunscreen use one week later, beyond self-reported attitudes and behavioral intentions (Falk et al., 2010). Building on this evidence, the current study evaluates the neural basis of an intervention that combines gain-framed health messaging and self-affirmation to reduce sedentary behavior in older adults. Specifically, we examined baseline activity during a novel fMRI paradign in VS and vmPFC, regions known to play critical roles in reward valuation and health message processing, to better understand the neural mechanisms supporting combined gain framed messaging and self-affirmation in older adults.

Technological tools, such as fitness trackers and smartphone applications, offer promising solutions for reducing sedentary behavior, particularly when integrated with evidence-based behavioral strategies (Sullivan & Lachman, 2017; Burke et al., 2011). The COVID-19 pandemic accelerated the adoption of remote interventions, enabling older adults to address barriers like limited mobility and restricted access to resources (Frost et al., 2020). However, the efficacy and feasibility of remotely delivered interventions remain understudied, particularly among individuals with low socioeconomic status or limited digital literacy. In the current study, we tested the efficacy of combined daily gain-framed health messages with self-affirmation in a technology-based randomized controlled trial that aimed to reduce sedentary behavior in older adults. Additionally, informed by principles of implementation science, this study employs a hybrid type 1 efficacy and implementation design to evaluate the adoption, implementation, and sustainability of this intervention (Proctor et al., 2013). Insights gained from this study may inform scalable, accessible solutions to support active aging in vulnerable populations. The overarching goals of this work are to test a novel strategy to reduce sedentary behavior in older adults and to characterize the extent to which individual differences in reward processing predict behavioral change.

## Methods

### Participants

We recruited 48 sedentary, cognitively healthy older adults from the greater Boston area at the outbreak of the COVID-19 pandemic. There were 8 participants who completed the initial consent visit but did not begin the study (e.g., dropped out because of change of eligibility and delayed start due to in-person data collection restrictions) while 40 participants were enrolled and began the behavioral randomized controlled trial and 38 (95%) participants completed the study. Due to pandemic restrictions, MRI session participation was made optional, and 14 participants opted to complete this part of the baseline assessment. Participants were recruited through Facebook advertisement and flyers posted in the Boston area at Seniors Centres, bus stops and multi-faith organizations. Potential participants underwent an initial phone screening interview for the following inclusion criteria: (1) Engagement in less than 150 minutes of moderate-to-vigorous physical activity (MVPA) per week or more than 8 hours of sitting time per day; (2) cognitively unimpaired as determined by a score on the Telephone Interview of Cognitive Status (TICS) >=26 (Fong et al., 2009); (3) access to a computer or smartphone with internet; (4) able to walk without experiencing severe pain; (5) no diagnosis of major psychiatric or neurological illness. For more details on inclusion and exclusion criteria, please refer to the protocol Ai et al. (2021).

## Measurements

### Questionnaires

Demographic information in all participants collected at the baseline visit included age, sex, ethnicity and education level. To address our hypothesis that reward and motivation-related individual differences predict behavior change, we acquired the following motivational and reward-related inventories: Temporal Experiences of Pleasure Scale (Gard et al., 2006) and the Apathy Evaluation Scale (Marin et al., 1991). To assess the extent to which potential barriers including comfort with technology and socioeconomic status influence intervention adherence, participants completed the Mobile Device Proficiency Questionnaire (Roque & Boot., 2018) and we acquired information about particpants’ socioeconomic status. In addition to the primary outcome (i.e., sedentary time), mental health status was considered as an exploratory outcome. A description of questionnaires in the current study is included in Table 1. During the last visit of this study, all participants completed feasibility questions such as “How feasible did you find completing the 6-week intervention?” and collected their answer on a Likert-scale from 1 (Not feasible at all) to 10 (Very feasible).

**Table 1.** Description of the inventories administered to participants in the study.

| Measure | Timepoint | Description |
| --- | --- | --- |
| Temporal experience of Pleasure Scale | Pre and Post intervention | This reward-related scale was developed and validated to assess individuals' ability to experience pleasure (Gard et al., 2006). The scale contains 18 questions with the subdimensions of anticipatory and consummatory pleasure. |
| Apathy Evaluation Scale | Pre and Post intervention | This 18-item scale was developed and validated to assess the degree of motivational impairment across cognitive, emotional and behavioral domains (Marin et al., 1991). The total score ranges from 18 to 72, with higher scores indicating more apathy. |
| Mobile Device Proficiency Questionnaire (MDPQ-16) | Pre intervention | This 16-item scale was developed and validated to assess individuals' ability to use and perform tasks with a mobile device (Roque & Boot, 2018). |
| Geriatric Depression Scale | Pre and Post intervention | This widely used and validated 15-item questionnaire was developed to assess depression symptoms in older adults with and without neurodegenerative disease (Yesavage et al., 1982). Greater scores suggest depression |
| Geriatric Anxiety Inventory | Pre and Post intervention | This validated 20-item questionnaire was developed to assess anxiety symptoms in cognitively unimpaired and impaired older adults (Pachana et al., 2007) |

**Table 2.** Baseline descriptive information for intervention and active control groups.

| Baseline variables | Intervention group<br>(n=21) | Active control group<br>(n=19) |
| --- | --- | --- |
| Age (Mean ± SD) | 70.0 ± 5.4 | 68.4 ± 5.0 |
| Sex | Female = 15, Male = 6 | Female = 18, Male = 1 |
| Race | Black/African American=7,<br>White/Caucasian=12,<br>Asian/Pacific Islander=2, | White/Caucasian=17,<br>Asian/Pacific Islander=1,<br>Other=1 |
| Years of formal education<br>(Mean ± SD) | 16.6 ± 3.6 | 16.0 ± 3.6 |
| Income (US \$) | < \$20,000=6,<br>\$20,000-\$34,999=6,<br>\$35,000-\$49,999=2,<br>\$50,000-\$74,999=1,<br>> \$75,000=4<br>Not specified=2 | < \$20,000=0,<br>\$20,000-\$34,999=3,<br>\$35,000-\$49,999=5,<br>\$50,000-\$74,999=4,<br>> \$75,000=5<br>Not specified=2 |
| Sedentary time at baseline<br>(ActivePAL; Mean ± SD) | 8.78±1.89 hours/day (n=19) | 7.83±2.46 hours/day (n=16) |

### Sedentary time and physical activity monitoring

The primary outcome of this intervention was time spent sedentary, which was measured with an activPAL inclinometer (activPAL, PAL Technologies, Glasgow, UK). The activPAL was attached to the mid-anterior surface of the right thigh. Physical activity (secondary outcome) was measured using GT9X Link accelerometers (ActiGraph LLC, Pensacola, FL) as a secondary outcome. The GT9X was worn on the non-dominant wrist. Participants were instructed to wear both activPAL and ActiGraph 24 hours per day for continuous seven days at the baseline (week 0), midpoint (week 3), and endpoint (week 6) of the intervention, and were mailed to participants. For data cleaning, ActiGraph data were included in the analysis if participants had a minimum of four valid wear days (Trost et al., 2005), while activPAL data were included if participants had a minimum of five valid wear days (Aguilar-Farias et al., 2019). Participants were also asked to fill out a sleep log, where they reported the time they went to bed and got out of bed each day. Additionally, they also noted the time when they removed and replaced either device each day during the assessment week. These information were used to remove sleep time and non-wear time out of daily movement recordings in later processing. Prior to each of the assessment weeks, there was a training visit and opportunities for participants to ask questions during a conference call to ensure that they correct device use.

### Intervention

Participants were randomized into the intervention or the active control groups through the REDCap randomization function, before the baseline assessment and MRI session, that was administered through the Harvard Catalyst Biostatistical Group. The intervention was 6-weeks, during which participants received daily messages via email or text, depending on their preference. The daily messages were launched via OTree, an open-source platform (Ai et al., 2021). Unique links were assigned to each participant at enrollment. The intervention combined gain-framed messages and self-affirmation practice. Prior to the intervention, participants ranked eight core values based their importance (i.e., family and friends, politics, religion, creativity, money, independence, humor, spontaneity). The list of values were chosen based on a previous study (Falk et al., 2015). Participants in the intervention group received daily prompts to reflect on their highest-ranked core value (e.g., ‘Think of a time when family and friends inspire you’). This was followed by a daily gain-framed health message about physical activity (e.g., ‘As you move around more, your body can use blood sugar. This can keep your arteries healthy.’) (Fig 1b).

**Figure 1.**
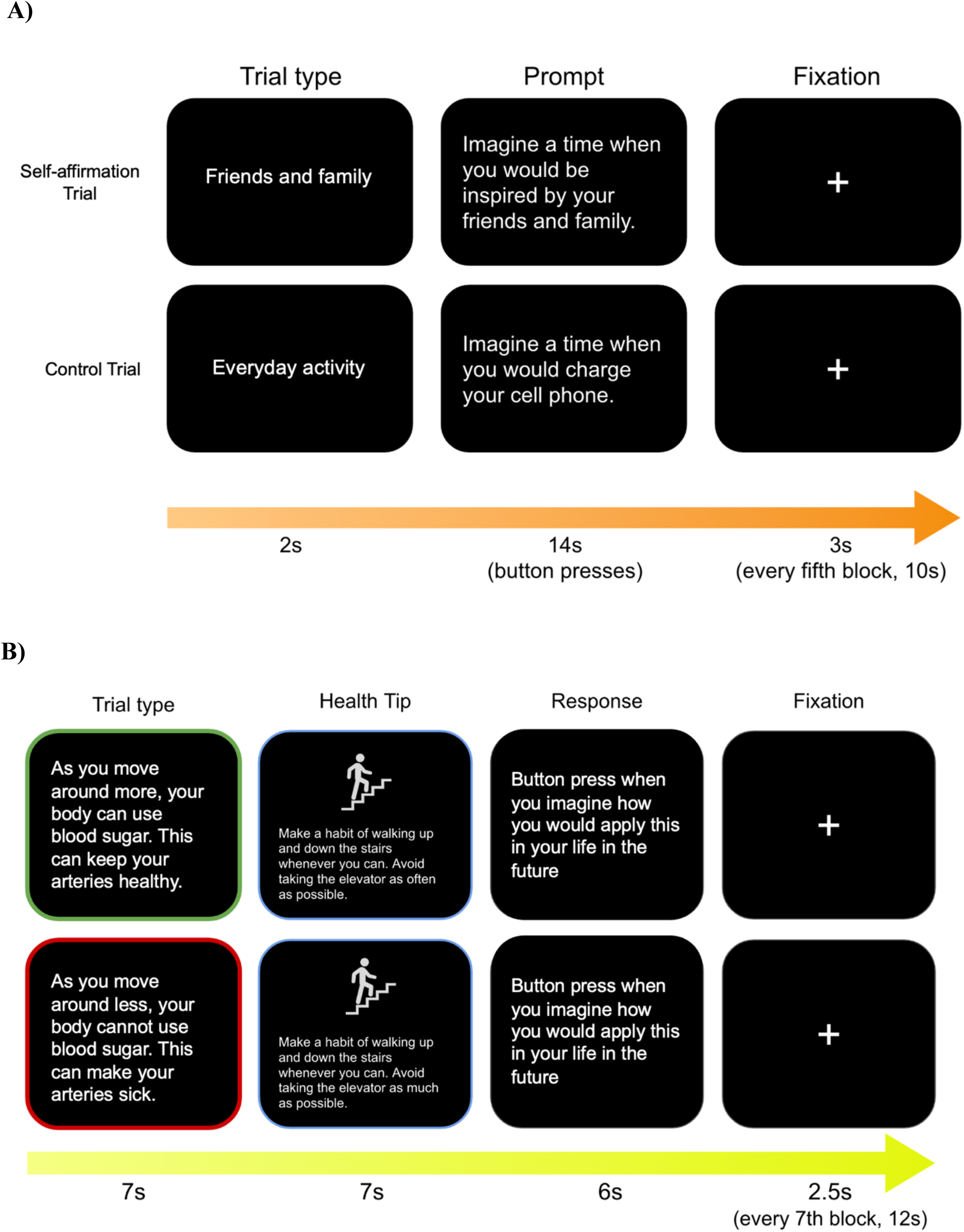
Example trials for the self-affirmation task (A) and the health-message task (B) as presented to participants. Gain-framed trials are highlighted in green (shown to the intervention group), and loss-framed trials are highlighted in red (viewed by the active control group).

### Control

The active control group received the same number of daily prompts as the intervention group. However, the self-affirmation messages encouraged participantes to reflect on an everyday activity (e.g., think of a time when you charge your phone) and the health messages had the same topic as the intervention group but in a loss-framed way (e.g., ‘As you move around less, your body cannot use blood sugar. This can make your arteries sick.’). Health messages focused on reducing sedentary behaviour. These daily self-affirmation and health messages consisted of a combination of the same messages used in the fMRI tasks and novel messages. Health messages were chosen based on a previous study (Northoff and Carstensen, 2015). The number of times participants completed the survey was recorded to indicate the overall intervention adherence. In both groups, adherence was defined as the number of days the participants logged on to the OTree daily messaging technology platform.

### Behavioral data analyses

Behavioral data were checked for completeness and accuracy for missing or invalid data. Baseline demographic information and sedentary time were tested for potential group differences. Mixed effect models (i.e., within-subject effect: pre- and post-intervention * between-subject effect: intervention and active control group) were estimated to investigate both primary outcome changes (i.e., sedentary time) and secondary outcome changes (i.e., physical activity engagement, depression and anxiety) across the intervention compared to the active control using lm4 package. Covariates including age and sex, and individual random intercept effect were included in the model. Multiple linear regression models were estimated to investigate whether individual differences in apathy level and reward sensitivity predict sedentary time change.

### Functional magnetic resonance imaging (fMRI) data acquisition

There were 14 participants who opted to perform MRI scans and completed the baseline neuroimaging acquisition. All baseline MRI data were acquired before the start of the intervention. Nine participants were randomized into intervention group, while 6 participants were in the active control group. Participants underwent scanning in a 3 Tesla Siemens PRISMA scanner with a 64-channel head coil. T1-weighted (T1w) images were obtained with a high-resolution three-dimensional (3D) Magnetization-Prepared Rapid Acquisition with Gradient Echo (MPRAGE) sequence (Sagittal, 0.8mm isotropic resolution, TE/TI/TR=2.31/1000/2500ms, Field of View (FOV) 256mm, 208 slices). FMRI task-based data were acquired with Echo Planar Imaging (EPI) sequences. Each task was divided into two separate runs with same time length, in order to provide an opportunity for participants to rest (Resolution: 2*2*2mm, TE/TR=37/800ms, Multiband factor=8, 72 slices, 534 or 627 measurements for each run).

### FMRI task-based paradigms

#### The Self Affirmation Task

After randomization, participants completed an event-related fMRI task at the outset of the study. This task contained 42 trials in total and included 21 value trials as the self-affirmation manipulation, where the intervention group was instructed to reflect on their highest-ranked core value, while the active control group was instructed to reflect on their lowest-ranked core value. As a within-subject control, there were 21 control trials, where both groups were instructed to reflect on neutral statements about their everyday activity. In each value trial, a cue (i.e., either value or daily activity) was displayed for 2 seconds, and followed by the value prompt (e.g., imagine a time when you would be inspired by your family and friends) for 14 seconds. During the prompt, participants were asked to make a button press whenever they thought of an example for the prompts. An example of task trials was displayed in Figure 1a.

#### The Health Messaging Task

Following the self-affirmation task, participants performed the health-messaging. This task contained 42 trials in total (Figure 1b). The intervention group received gain-framed health messages while the active control group received loss-framed health messages. Each message was followed by a health tip, the content of which was identical across groups. As a within-subject design, there were 21 health message trials and 21 daily activity control trials for both groups. During each trial, participants viewed the health-related message or control messages about daily activities for 7 seconds, and a subsequent corresponding tip for 13 seconds. After the tip, participants were given 6 seconds and asked to press the button whenever they imaged how they might apply this tip in their life. An example of a task trial is displayed in figure 1b. Both tasks were programmed in PsychoPy software (Peirce, 2007) and were also administered for participants who opted out MRI sessions in a videoconferenced call using Pavlovia (https://pavlovia.org/) so that all participants completed these computerized paradigms at baseline.

### FMRI Preprocessing and Statistical Analyses

Quality control of fMRI data, including naming standards, correct number of volumes per sequence and correct sequence parameters, was done following data collection. Data were converted to NIFTI format and organized in Brain Imaging Data Structure (BIDS) format. High-resolution anatomical images of the brain were extracted using the Brain Extraction Tool (Smith, 2002) and subsequently normalized to a standard Montreal Neurological Institute (MNI) coordinate space. The fMRI data preprocessing and statistical analyses were performed with FEAT in FSL (version 6.0.0). Motion correction, smoothing with a 6-mm full-width at half-maximum Gaussian Kernel, and high-pass filtering of 100 seconds applied to the functional data. Effects of large motion outliers within the time series were detected and regressed out via the FSL motion outlier software (https://fsl.fmrib.ox.ac.uk/fsl/fslwiki/FSLMotionOutliers). All functional images were registered to high-resolution T1 anatomical images and normalized to MNI space. Both the self-affirmation task and health messaging task were modelled using double gamma convolution to fit the evoked hemodynamic response.

For the first level analysis, parameter estimates of contrasts were created to quantify activation underlying self-affirming and gain-framed messaging (i.e., self-affirmation prompt > daily activity prompt) in the self-affirmation task, and activation underlying health message viewing (i.e., framed health message > daily activity message), and health tips viewing (i.e., health tips > daily activity tips) in the health-message task. For the second level analysis of both tasks, parameter estimate maps originating from the first level analysis were carried forward to examine group-level differences between the intervention group and the active control group.

These were estimated with FSL’s Local Analysis of Mixed Effects (FLAME) with a cluster probability threshold of p<0.05 and voxel-wise threshold of z>2.34. We adopted a hypothesis-driven approach by selecting prior-defined Regions of Interest (ROIs) for higher level analysis. Specifically, the masks of ventromedial prefrontal and ventral striatum, which were previously proved to associate with subjective value (Bartra et al., 2013), were applied as ROIs (Figure 2), in order to investigate group difference in activation of regions involved in subjective valuation.

**Figure 2.**
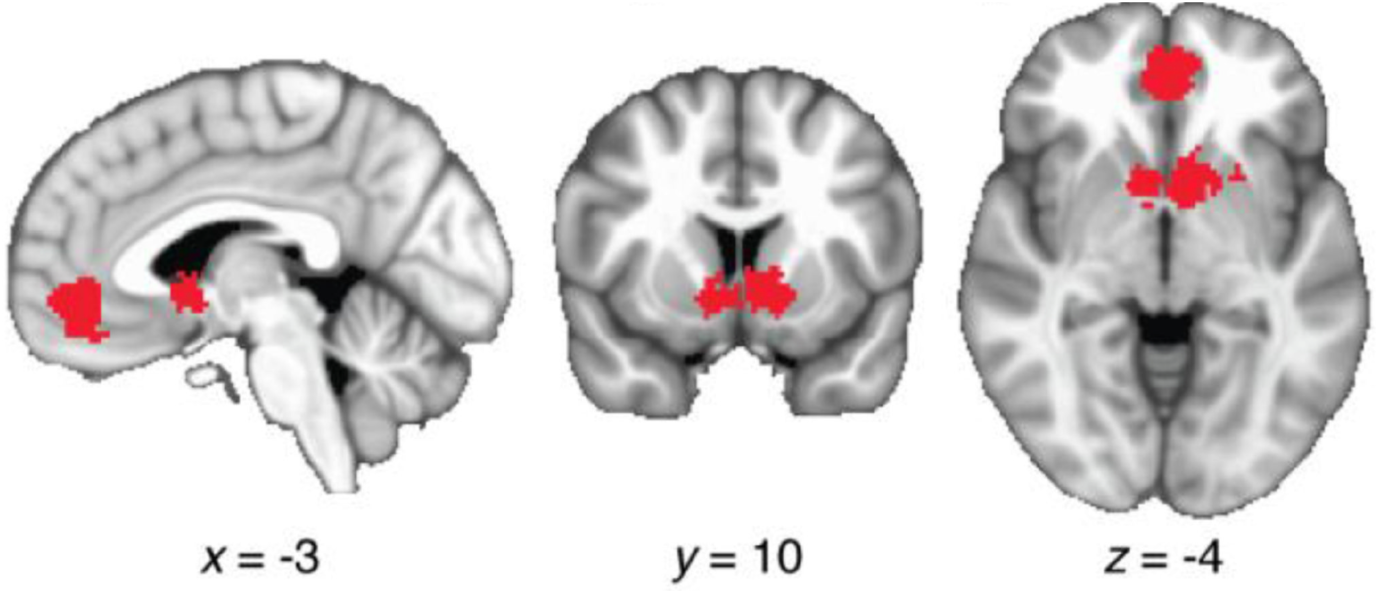
Regions of Interest in the reward valuation network. These were functionally defined based on a meta-analysis (Bartra et al., 2023) within ventromedial prefrontal cortex and ventral striatum.

## Results

### Descriptive information

There were no group differences between the intervention and the active control groups for demographic information or baseline sedentary time for the 40 participants. At baseline, there were five participants who did not have sufficient valid days of sedentary time data, therefore, baseline ActivPAL data from a sample of 35 participants are displayed in the table below.

### Behavioral results

For the daily messaging OTree technology platform use, participants showed an overall high intervention adherence (0.92±0.08; figure 3a), that did not differ by group (t=-1.2, *p*=0.23; figure 3b). During the final debriefing visit, participants rated the feasibility of implementation of the intervention in their daily lives on a likert-scale from 1 to 10 highly overall (8.75±1.73 for intervention group, 9.77±0.60 for active control group), across both groups (Figure 3c). There was no group difference in mobile technology proficiency (67.95±11.11 for active group, 70.47±9.64 for active control group; t=-0.46, *p*=0.65; Figure 3d). No significant relationship was found between either socioeconomic status or mobile technology proficiency and intervention adherence.

**Figure 3.**
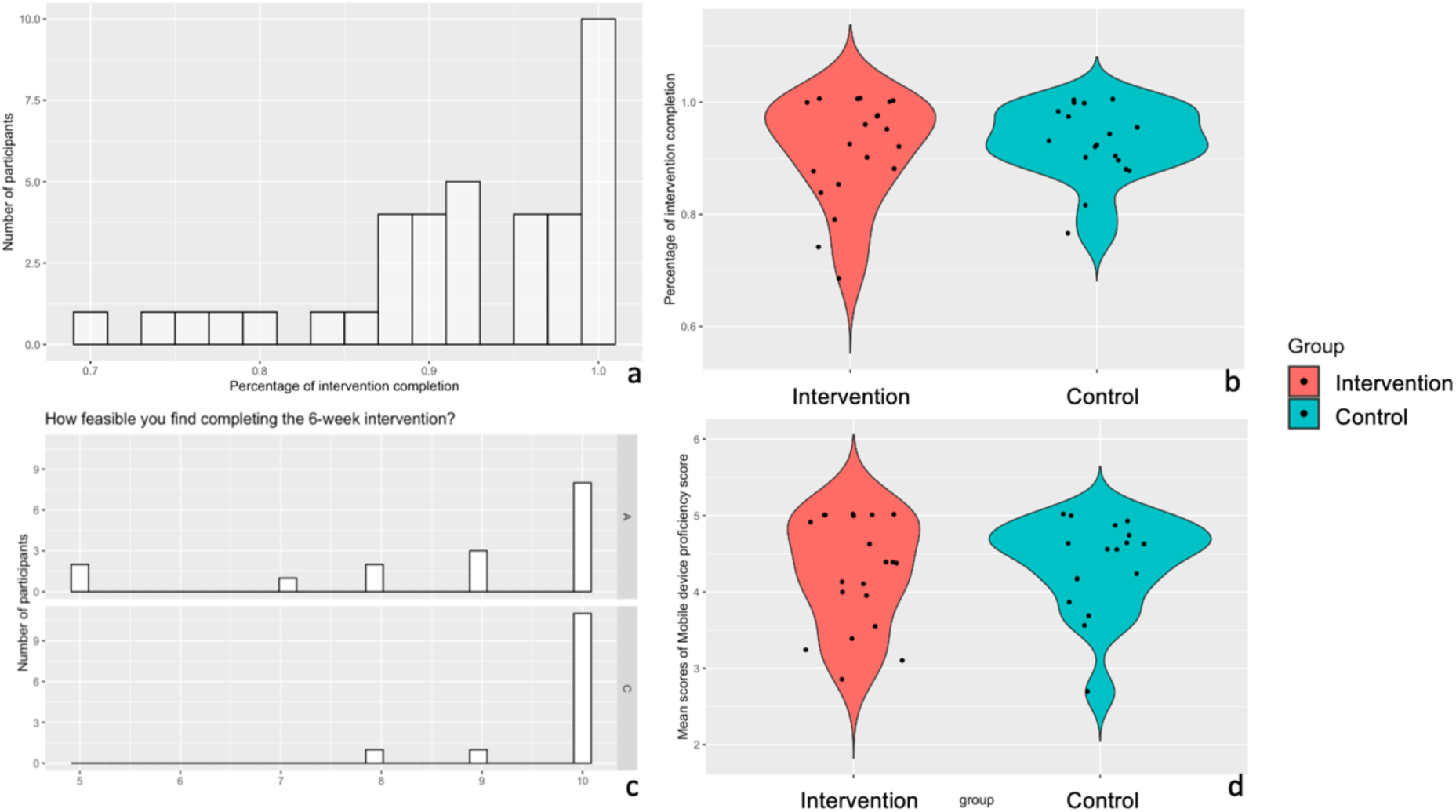
Overall daily survey compliance (left) and overall compliance distribution by group (right).

After removing participants who did not have enough valid days either for baseline or post-intervention assessments for either ActiGraph or ActivPAL data, there were 27 participants in total for the sedentary time (i.e., ActivePAL) analysis, and 29 participants in total for the MVPA (i.e., ActiGraph) analysis. For the primary outcome, there were no group differences in sedentary time changes after the intervention (β=-0.25, t=-0.48, p=0.63; Figure 4). For secondary outcomes, there were no group differences in MVPA (β=0.29, t=0.82, p=0.42), depression (β=0.11, t=0.44, p=0.66), or anxiety (β=-0.11, t=-0.47, p=0.64) changes after the intervention compared to baseline (Figure 4).

**Figure 4.**
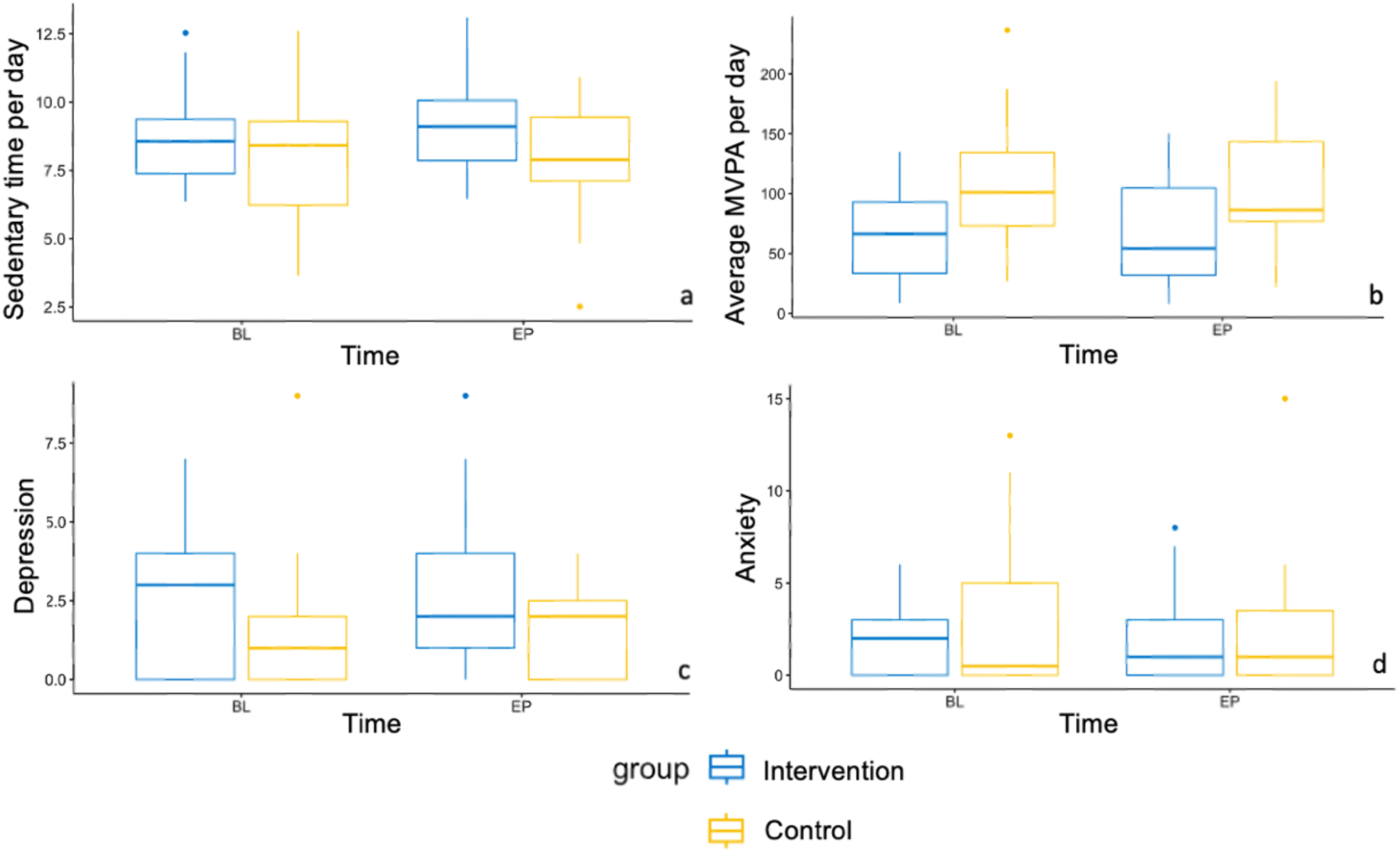
Boxplots comparing sedentary time (a), moderate-to-vigorous physical activity (MVPA) (b), depression (c), and anxiety (d) at baseline (BL) and post-intervention (EP), and, between intervention and active control groups.

We conducted an exploratory analysis to determine whether individual differences in reward and motivational traits predict the degree of sedentary behaviour change after the intervention compared to baseline across all participants, regression analyses were performed. Increased anticipatory pleasure, an index of reward sensitivity at baseline, showed a significant association with greater reduction in sedentary time after the intervention compared to baseline (r^2^=0.303, p=0.005; Figure 5). However, there was no significant association between level of consummatory pleasure (r^2^=0.003, p=0.865), or apathy level (r^2^=0.001, p=0.782) and sedentary behavioral change. Age, sex, and baseline sedentary time were controlled as covariates of non-interest across all analyses.

**Figure 5.**
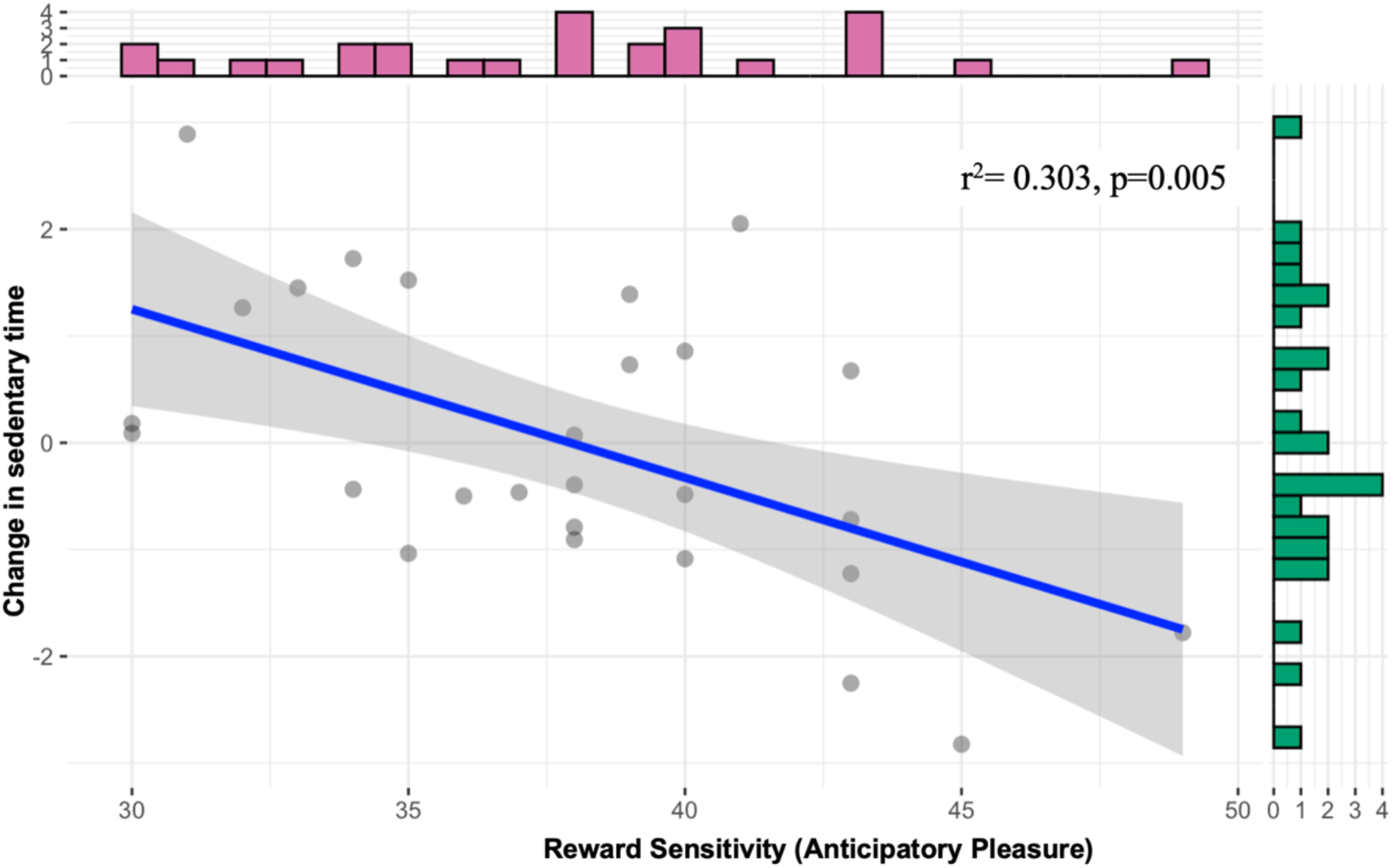
The relationship between reward sensitivity (anticipatory pleasure) and changes in sedentary time in all participants after the intervention period, compared to baseline.

### FMRI task results

For the health messaging task, the intervention group showed greater activation in left VS under the contrast of health tips > daily activity tips compared to the active control group (figure 3; number of voxels=93, coordinate -12, 14, 2). However, our analysis did not show any group difference in activation of VS or vmPFC under contrast of self-affirmation prompt > daily activity prompt of the self-affirmation task, or under contrast of framed health message > daily activity message of the health messaging task.

**Figure 3.**
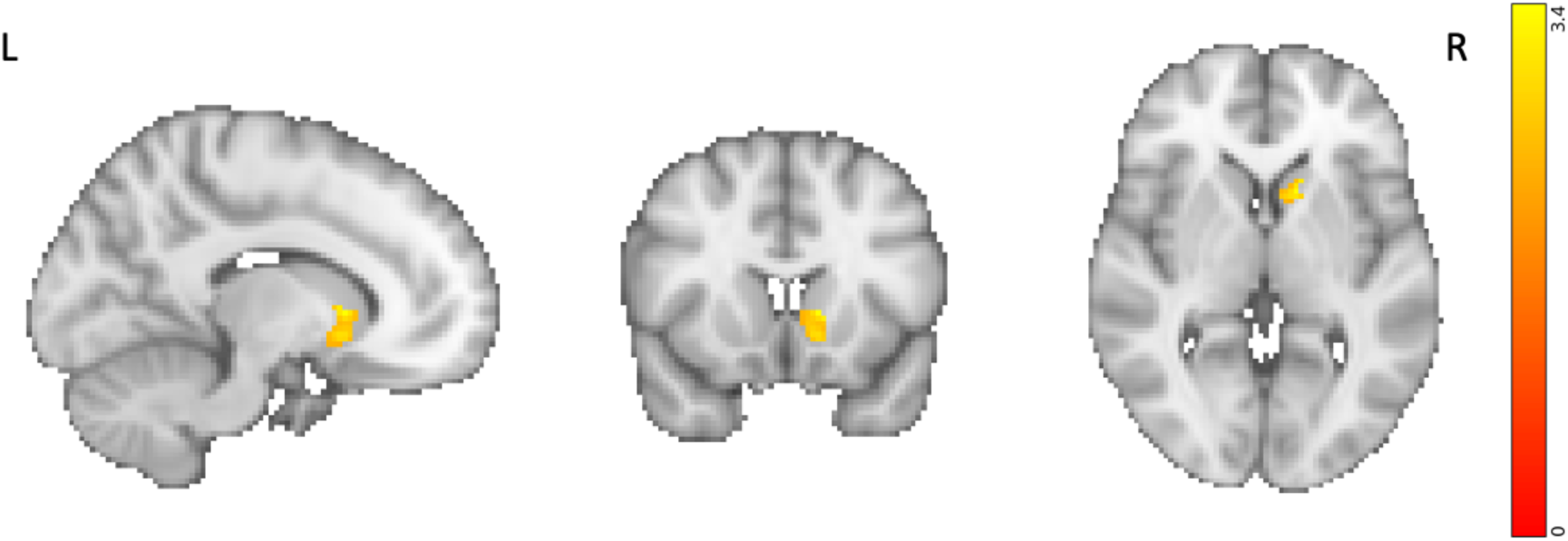
A significant cluster within the left ventral striatum showed greater activation in participants in the intervention, compared to the active control, group at baseline under the contrast of health tips > daily activity tips. Univariate activation analysis results are overlaid on the MNI template brain and corrected for multiple comparisons.

## Discussion

The goals of this study were to (1) evaluate the efficacy of combined self-affirmation and gain-framed health messaging in a novel intervention aimed at reducing sedentary behavior in older adults, (2) assess the feasibility of completing a remotely administered, technology-based intervention, and (3) determine whether baseline individual motivational differences predict changes in sedentary behavior. Additionally, as an exploratory aim, this study examined the neural basis of combined gain-framed health messaging and self-affirmation in older adults during the viewing of potentially threatening health messages. While no significant differences in sedentary behavior change were observed between groups, the findings indicated that this technology-based intervention was highly feasible for older adults. In particular, no significant relationship was found between socioeconomic status or mobile device proficiency and adherence to the intervention. Furthermore, the results revealed that individual differences in reward sensitivity at baseline predicted the change in time spent sedentary. Finally, during a task-based fMRI paradigm at the outset of the study, participants in the intervention group exhibited heightened activity in the left ventral striatum, a node within the reward-network, compared to the active control group when viewing health information relative to everyday information.

The intervention’s feasibility was supported by a high adherence rate, with older adults completing an average of 92% of the intervention. In addition, participants reported high subjective feasibility of implementing this intervention in daily life. This finding is particularly important because older adults often encounter greater barriers to adopting health applications compared to younger adults, including reduced perceptual, cognitive, and motor abilities, lower familiarity with technology, and a lack of support when using new technologies (Li et al., 2021; van Acker et al., 2023). Previous mobile health interventions have shown promise in promoting physical activity among older adults; however, user engagement with these interventions was often low and inconsistent (van Acker et al., 2023). Specifically, older adults are less likely to adopt technology if they do not recognize its benefits, or if it is perceived as time-consuming, unreliable, or burdensome (Matthew-Maich et al., 2016; Rogers & Fisk, 2010). Implementation science indicates that the factors influencing an intervention’s implementation differ from those that determine its efficacy, making it essential to identify both facilitators and barriers to integrating interventions into individuals’ daily lives (Bauer & Kirchner, 2020). Nevertheless, there is currently limited research on best practices for implementing health applications in the daily lives of older adults (Matthew-Maich et al., 2016). Thus, the results of this study provide promising evidence for the ease of implementing a remotely administered, technology-based intervention into the daily routines of older adults.

This study examined two common barriers to intervention adherence among older adults: lower socioeconomic status and mobile device proficiency. Prior research has shown that older adults with lower socioeconomic status are less likely to adopt health technologies compared to those with higher socioeconomic status (Chan et al., 2023). This disparity is attributed to lower income groups having reduced access to technologies, and less experience and education that facilitate their use (Darrat et al., 2021). Additionally, low device proficiency has consistently been associated with diminished adoption of health technologies among older adults likely because technology proficiency directly influences older adults’ perceived ease of use (Fowe & Boot, 2022; Mitzner et al., 2019; Hawley-Hague et al., 2014). To enhance older adults’ engagement and adherence, it has been suggested that health applications should be simple and tailored to users’ needs (van Acker et al., 2023). Notably, this study found no relationship between socioeconomic status or mobile device proficiency and intervention adherence, indicating that older adults could effectively engage with the technology over time. This finding is important, as consistent and sustained engagement with health applications is a promising strategy for achieving effective and lasting behavior change (Bakker & Rickard, 2018; Kowalski et al., 2024).

In addition to demonstrating high feasibility, the results of the intervention revealed that individual differences in reward sensitivity predicted reductions in sedentary behavior across both groups. Specifically, greater anticipatory pleasure was associated with larger reductions in time spent sedentary, while no significant relationship was found between consummatory pleasure and sedentary time. This finding aligns with prior research indicating that reductions in sedentary behavior are reinforced by the anticipated pleasure associated with an active lifestyle, including enhanced exploration and social interactions (Meredith et al., 2023; Phoenix & Orr, 2014). Anticipatory and consummatory pleasure can be differentiated into the ‘wanting’ and ‘liking’ aspects of motivation, respectively (Selby et al., 2020). At the neural level, anticipatory pleasure is associated with activity of the dopaminergic reward system, including the ventral striatum, while consummatory pleasure is driven by the endogenous opioid system (Nguyen et al., 2021; Hoflich et al., 2019) and implicates the orbitofrontal cortex, a brain region involved in sensory integration that represents subjective, hedonic experience (Kringelbach, 2005). Previous studies have shown that activity of the dopaminergic reward system, including the vmPFC and VS, predicts reductions in sedentary behavior in younger adults (Cascio et al., 2016; Kang et al., 2018). Further, anticipatory pleasure has been shown to predict higher physical effort expenditure for rewards (Geaney et al., 2015). Frameworks of effort-based decision making suggest that engaging in effortful activities involves evaluating whether the value of a reward outweighs the cost of expending effort (Shenhav et al., 2013). The theory of effort minimization in physical activity posits that sedentary behavior arises from people’s automatic tendency to conserve energy by minimizing effort expenditure (TEMPA; Cheval & Boisgontier, 2021). To decrease sedentary behavior, the theory suggests that both controlled (e.g., reasoned attitudes, intentions) and automatic (e.g., affective reactions) processes must be sufficiently strong to override the innate tendency to minimize effort. Experimental research supports this model, demonstrating that sedentary behavior is automatic and even rewarding for adults (Cheval et al., 2018; Falck et al., 2024; Morris et al., 2022).

Contrary to our hypothesis, there were no significant differences in sedentary behavior change between older adults who completed a daily self-affirmation task followed by a gain-framed health message compared to a well-matched active control group that reflected on an everyday activity followed by a loss-framed health message. Prior research that examined self-affirmation interventions on physical activity engagement and sedentary behavior in younger adults is mixed. In support of its benefit, prior behavioral and neuroimaging studies in younger adults have shown that reflecting on core values reduces sedentary behavior and increases physical activity (Cascio et al., 2016; Falk et al., 2015). In contrast, a large behavioral study examined the influence of value-affirmation on gym visits in 1628 adults (mean age = 30.8 years) and showed no group difference between intervention and active control participants (Milkman et al., 2021). There is also support for the potential benefit of self-affirmation across multiple behavioral domains other than physical activity and sedentary behavior in younger adults. For example, self affirmation was shown to support successful educational achievement (Miyake et al., 2010; Cohen et al., 2009) reduced alcohol consumption (Harris & Napper, 2005), smoking cessation (Harris, Mayle, Mabbott, & Napper, 2007), and healthy dietary choices (Schuz et al., 2017). While meta-analyses of self-affirmation interventions have shown overall positive effects in younger adults, these effects tend to be small-to-medium and variable (Epton et al., 2015; Sweeney & Moyer, 2015), with individual-level factors such as risk level and self-esteem impacting successful behavior change (Schuz et al., 2017). This variability suggests that the efficacy of self-affirmation interventions in younger adults may depend on the specific contextual factors in which they are introduced. For example, they may be most effective for individuals who are under psychological threat that inhibits their openness to behavior change or in contexts where resources for change are available (Cohen & Sherman, 2014; Borman et al., 2018; Ferrer & Cohen, 2018). Prior meta-analyses have also highlighted the need to test self-affirmation as a strategy to enhance health behavior engagement in older adults (Sweeney & Moyer, 2015). Only one other behavioral trial tested the influence of self-affirmation on behavior change in aging: A randomized controlled trial in 960 older adults showed that self-affirmation did not influence medication adherence among diverse older adults (Daugherty et al., 2021). In older adults, self-affirmation may relieve negative emotions (Dang et al., 2023), however, there is currently no evidence supporting its impact on behavior in this population.

Ferrer and Cohen (2018) proposed a framework outlining the contextual factors that influence the effectiveness of self-affirmation for behavior change. Their framework posits that for self-affirmation to be effective, three conditions must be met: 1) the presence of a psychological threat to the self that hinders behavior change, 2) the availability of resources (e.g., training, support, guidance) to facilitate behavior change, and 3) the temporal proximity of self-affirmation to the perceived threat and resources for change. Generally, older adulthood is characterized by enhanced psychological well-being and emotion regulation skills compared to younger adulthood (Riediger & Bellingtier, 2022). Moreover, older adults often hold positive attitudes toward exercise (Hurst et al., 2023; Jadczak et al., 2018) and health messaging (Lam et al., 2014). A reason for the discrepancy in the efficacy between younger and older populations may be attributed to changes in differing levels of perceived psychological threat. Given these insights, future studies examining the impact of self-affirmation on behavior change in older adults should include measures of perceived threat at baseline to identify features of older adults that might benefit most from reflecting on core values. Additionally, future research could enhance effectiveness by providing resources to facilitate behavior change in close temporal proximity to the perceived threat.

This study examined the impact of a combined self-affirmation manipulation with health message framing on changes in sedentary behavior. Previous research has indicated that gain-framed health messages are generally more effective at promoting physical activity among older adults compared to loss-framed health messages (Liu et al., 2019; Notthoff & Carstensen, 2014), in contrast to the findings in the current study. One study conducted in individuals with spinal cord injury found loss-framed messges were more effective for eliciting cognitive processing and changing leisure time physical activity beliefs and intentions (Bassett-Gunter et al., 2013). Nonetheless, there are instances where both types of message framing have been shown to be equally effective in encouraging behavior change. For example, prior studies have demonstrated that the effectiveness of message framing on health behaviors may not be significant unless individual differences in motivation and perceived risk are taken into account. For instance, gain-framed messages have been found to be particularly effective at increasing physical activity among individuals with low levels of walking self-efficacy, grit, and consideration of future consequences (Jensen et al., 2018), as well as among those who perceive a high level of risk in not engaging in the behavior (Apanovitch et al., 2003). It is possible that the participants in this study exhibited distinct characteristics regarding these moderators, leading to comparable effects of both types of health message framing message on sedentary behavior change. Therefore, future research should examine how moderators such as grit, consideration of future consequences and degree of risk perception to better understand their influence on behavior change in older adults, for example by applying the operating conditions framework (Rothman & Sheeran, 2021).

The fMRI results from the health messaging task indicated that the intervention group demonstrated increased activity in the left VS when presented with health tips compared to daily activity tips, relative to the active control group following self-affirmation. The health tips were identical in both groups, but were preceded by gain-framed health messages in the intervention group, whereas the active control group received loss-framed messages prior to viewing the health tips. The ventral striatum is a subcortical region within the reward network, recognized for its role in the affective processing of rewards and its activation in response to positive emotional stimuli (Pool et al., 2022; Wager et al., 2008). These findings suggest that VS activity is heightened when viewing health tips that follow gain compared to loss message framing, possibly because gain-framed health messages enhance the rewarding properties of subsequent health information for older adults. This outcome aligns with the positivity effect observed in older adulthood, where individuals exhibit a preference for positive over negative information during cognitive processing (Carstensen & DeLiema, 2018). However, combined with our behavioral findings, recruitment of the reward network was insufficient to effectively change sedentary behavior. This is in contrast to prior behavioral research showing that positive affect may serve as a mechanism linking the positivity effect to behavior change (Liu et al., 2019; Mikels et al., 2016). One study found that higher positive affect elicited by gain-framed messages contributed to an increased perception of the health message’s effectiveness among older adults (Liu et al., 2019). Additionally, Mikels et al. (2021) showed that gain-framed messages generated more positive affect than loss-framed messages, and that this positive affect predicted both the intention to enroll in and actual enrollment in a physical activity program for older adults. The activity of the VS may reflect the neural basis of positive affective evaluations of health information following positive message framing supporting behavior change. However, this study did not collect any self-reported data on affect elicited by the health messages which might explain the discrepant findings. Given the role of the ventral striatum in reward processing, these findings have implications for future behavior change strategies, suggesting that positive message framing may encourage favorable responses to health recommendations among older adults. Future studies are needed to investigate whether, and how, the neural response to health information following message framing relates to actual behavior change.

Despite the valuable insights gained from this study, some limitations must be addressed. First, the sample show recruitment bias towards overrepresentation of women and higher education, which raises concerns about the generalizability of the results across different sexes, education levels, and geographic regions. Additionally, the study was conducted at the onset of the COVID-19 pandemic, which may have influenced older adults’ ability to engage in physical activities (i.e., due to the closure of gyms, outdoor leisure areas, and social exercise groups), social isolation, and confidence in becoming more active. These restrictions introduced by the pandemic likely posed additional barriers to engaging in an active lifestyle, especially for older adults who experienced significant increases in sedentary behavior and diminished self-perceptions of physical abilities during the pandemic (Oliveira et al., 2022; Runacres et al., 2021). While the current results offer intriguing preliminary evidence regarding the neural basis of positive message framing in older adults, these findings should be replicated in future studies to enhance their validity given the limited sample size. Larger neuroimaging datasets are necessary to capture reliable and reproducible brain-behavioral phenotype associations (Marek et al., 2022).

In conclusion, this study demonstrates the feasibility of implementing a remotely administered, technology-based intervention among older adults. Our results point to a shift in influence and efficacy of self-affirmation on behavior change across the lifespan. We also found that while older adults showed recruitment of the reward network in the intervention group relative to the active control group at baseline, this did not translate into sedentary behavior change. Our results also point to the importance of reward-related processes in behavior change more generally, and showed an association between reward sensitivity and future sedentary behavior change across all participants. These findings critically inform future research aimed at enhancing older adults’ engagement in health behavior adherence.

## Data Availability

All data produced in the present study are available upon reasonable request to the authors

## Acknowledgements

This research was undertaken thanks in part to funding from a National Sciences and Engineering Research Council of Canada (NSERC) Discovery Grant (DGECR-2022-00299), an NSERC Early Career Researcher Supplement (RGPIN-2022-04496), a Fonds de Recherche Santé Québec (FRSQ) Salary Award, the Canada Brain Research Fund (CBRF), an innovative arrangement between the Government of Canada (through Health Canada) and Brain Canada Foundation, an Alzheimer Society Research Program (ASRP) New Investigator Grant, the Canadian Institutes of Health Research, the Canada First Research Excellence Fund, awarded through the Healthy Brains, Healthy Lives initiative at McGill University, and the National Institutes of Health (P30 AG048785) to MRG. Harvard Catalyst Biostatistics Consulting Service facilitated randomization via the Harvard Clinical and Translational Science Centre (National Centre for Research Resources and the National Centre for Advancing Translational Sciences (NCRR and the NCATS NIH, UL1 TR001102)) and financial contributions from Harvard University and its affiliated academic healthcare centers. This research was undertaken thanks in part to funding from Consortium pour l’Identification précoce de la Maladie d’Alzheimer-Québec (CIMA-Q) to NT and from the Canada First Research Excellence Fund and Fonds de recherche du Québec, awarded to the Healthy Brains, Healthy Lives inititative at McGill University to CSW.

